# Development of low-cost serological immunoassay to detect antiviral antibodies to Sars-Cov-2 Spike protein

**DOI:** 10.1101/2021.06.07.21258114

**Authors:** Luis Antonio Peroni, Jessica Toscaro, Camila Canateli, Gabriel Lima, Ana Pagliarone, Elaine Cardoso, Renata Viana, Jessica Bassani Borges, Hui Tzu Lin-Wang, Cely S. Abboud, Carlos Gun, Kleber Franchini, Marcio Bajgelman

## Abstract

Seroconversion to SARS-CoV-2 has been widely studied to evaluate infection spreading for epidemiological purpose, or even in studies of protective immunity in convalescent or vaccinated individuals. The viral particle has an envelope harboring the spike glycoprotein, which can be used as an antigen for assay development, to detect antiviral antibodies to SARS-CoV-2. Since several vaccines encode a spike subunit, the full length spike-based immunoassay should be a universal tool to evaluate seroconversion. In this manuscript, we propose a low-cost ELISA that can be used to detect antiviral IgG to SARS-CoV-2 in human serum.

## Introduction

Since COVID-19 outbreak, immune surveillance to SARS-CoV-2 has been extensively investigated by serological tests. In contrast to molecular assays such as qPCR that quantifies virus genome and has a narrow detection window, which spans about one week from symptoms onset, the serological assay may provide information from two weeks to a longer post illness period^1^. The basic principle of the serological immunoassay is based on a virus-specific antigen that mediates the capture of antiviral antibodies. Among virus antigens, stands out the SARS-CoV-2 spike glycoprotein^2–4^. The spike is the most exposed virus protein, decorates virus envelope and interacts with the cell surface receptor ACE2 which plays an important role mediating virus infection^5–7^. The spike-ACE2 interaction may be targeted by humoral immunity, which may prevent virus infection by neutralizing antibodies. These neutralizing antibodies have the ability to bind to a specific domain at spike protein blocking the interaction between the viral spike and the cell surface receptor ACE2. In this way, there are several investigations on passive immunization, using convalescent sera as a therapy for severe COVID-19 patients^8^. Some serological assays also use the spike binding domain (RBD) as a target antigen to detect neutralizing antibodies^9,10^. Besides humoral immunity, T cell response also has been investigated. Previous studies for SARS-CoV have demonstrated the presence of memory T Cells in ill-recovered patients, and these cells were able to respond to a rechallenge with a spike protein from SARS-Cov2 stimuli^11^. Most of recombinant vaccines that are being developed for COVID-19 use the full length or a spike domain as a target antigen^12^. The serological assays allow to observe dynamics of virus spreading, to explore protective immunity or even to verify seroconversion in vaccinated individuals^1,13,14^. Here in we describe a low-cost Enzyme Linked Immunosorbent Assay (Elisa) to detect antiviral antibodies, using a full length spike as target antigen.

## Results

### PCR positive SARS-CoV-2 individuals exhibit IgG anti-spike antibodies

In order to assess the assay selectivity, we chose 18 serum samples, collected from health professionals at the hospital Dante Pazzanese which were previously enrolled to PCR testing for COVID-19. Among these 18 samples, 9 samples were positively diagnosed to SARS-CoV-2 infection (Table 1).

**Table 1.** PCR testing for SARS-CoV-2 detection in enrolled individuals.

| sample | PCR |
| --- | --- |
| NS | NA |
| 1 | + |
| 2 | - |
| 3 | - |
| 4 | + |
| 5 | - |
| 6 | + |
| 7 | + |
| 8 | - |
| 9 | - |
| 10 | + |
| 11 | + |
| 12 | + |
| 13 | - |
| 14 | - |
| 15 | - |
| 16 | + |
| 17 | + |
| 18 | - |

The serum samples of these individuals were diluted 1:100 and loaded to Elisa plates previously coated with spike protein, to perform the anti-spike immunoassay. As seen in Figure 1, the SARS-CoV-2 PCR positive individuals showed a high signal of anti-spike IgG in the serological assay. The cut-off threshold was set as two-fold the signal of the negative control (NS).

**Figure 1.**
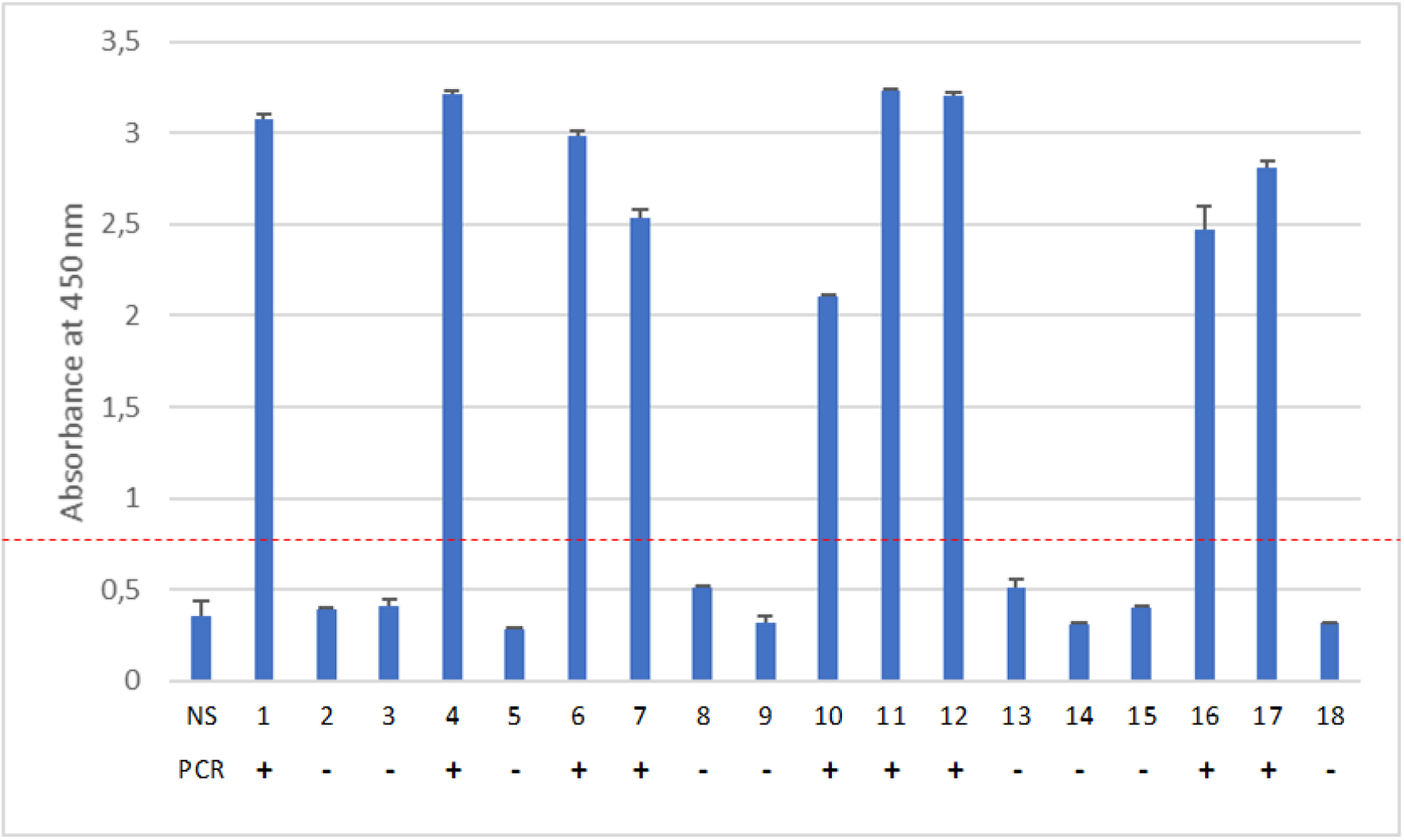
Immunoassay to detect anti-spike IgG antibodies from samples of PCR-diagnosed SARS-CoV-2 individuals. In this assay, serum samples were diluted 1:100 and loaded to SARS-CoV-2 spike-coated microplates. The antiviral spike-bound antibodies were detected with a secondary HRP-anti-human IgG antibody, following a reaction with TMB substrate and absorbance reading at 450nm. Below the X-axis, we indicate the sample number and the PCR result and NS = negative serum.

### Vaccinated individuals exhibited an increased signal of anti-spike IgG

The Elisa for anti-spike also may be used to evaluate the efficiency of vaccines. In this way, we collected and analyzed serum from healthy individuals that were given the CoronaVac vaccine. The CoronaVac vaccine is an inactivated SARS-Cov-2, that harbors spike protein^15^. The enrolled individuals are health professionals that work in the research department of the Brazilian Laboratory for Biosciences at the National Center for Research in Energy and Materials, and had no detectable level of anti-spike antibodies before vaccination. The serum was collected before vaccination, 15 days after the 1^st^ dose (1^st^ dose) and 15 days after the 2^nd^ dose (2^nd^ dose). As seen in Figure 2, there is an increase in antibody titer, as expected in vaccinated individuals, after the 1^st^ vaccine dose, which is boosted by the 2^nd^ dose.

**Figure 2.**
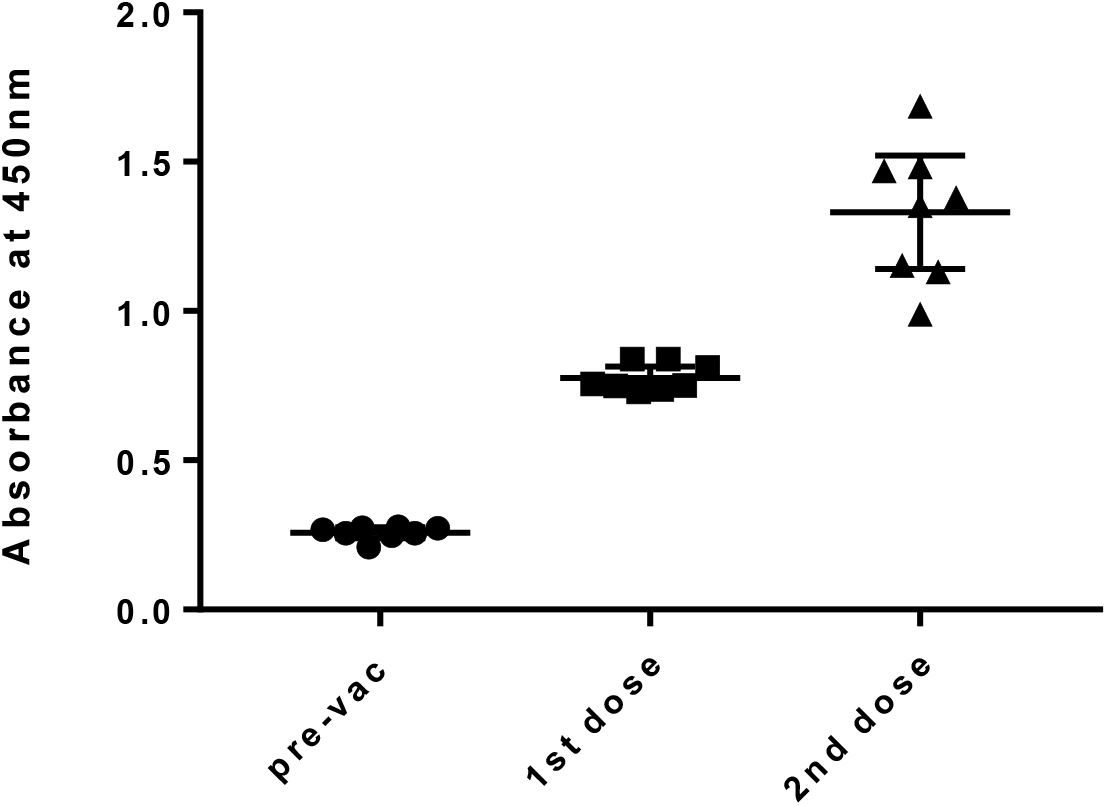
The Elisa assay may be used to monitor vaccine immunization. Serum samples were diluted 1:100 and loaded to 96-well plates previously coated with the spike antigen. Antiviral antibodies were detected with an HRP-anti human IgG. Pre-vac: serum collected before vaccination; 1^st^ dose: serum collected 15 days after the 1^st^ vaccine dose and 2^nd^ dose: serum collected 15 days after 2^nd^ vaccine dose.

## Conclusion

This serological testing employs a full length spike protein from SARS-CoV-2 as an antigen to capture human antiviral antibodies. The human antibodies that are bound to the spike-imobilized antigen are detected with a HRP-anti-human IgG antibody. In this way, we were able to confirm the presence of anti-viral IgG antibodies in the serum of SARS-CoV-2 individuals that were previously diagnosed by PCR, or even and increased anti-spike IgG signal in CoronaVac vaccinated individuals. The spike-based immunoassay showed high selectivity and may be adapted to detect other antibodies classes of clinical relevance, such as IgM. This same assay model can also be developed to detect other viral proteins, such as the Nucleocapsid protein. The Elisa immunoassay for detection of antiviral antibodies has potential for development of low-cost research assay or even diagnostic assay, providing a qualitative indication of reactivity for the SARS-CoV-2 Spike antigen.

## Material and methods

### Materials

| # | Item | Brand | Catalog |
| --- | --- | --- | --- |
| 1 | 96-well plate Maxisorp, Nunc | Thermo | 44-2404-21 |
| 2 | Sodium Carbonate, M = 105,99 g/mol | Sigma | 791768-500G |
| 3 | Sodium bicarbonate solution, M = 84,01 g/mol | Sigma | S5761-500G |
| 4 | Tween 20 | Sigma | P1379-250ML |
| 5 | 1X PBS | Thermo | 21600010 |
| 6 | TMB | Thermo | 002023 |
| 7 | Secondary HRP-Anti-human IgG | Sigma | A8792-2ML |
| 8 | BSA | Sigma | A7906-100G |
| 9 | 37% HCL | - | - |

### Preparation of buffers and reagents

Coating Buffer: 50mM Sodium Carbonate, pH 9.6

To prepare 1L, add in 800mL of H2O, 2.88g of sodium bicarbonate (84.01 g/mol) and 1.67 g of sodium carbonate (105.99 g/mol). Homogenize and make up to 1L with H2O. Check pH 9.6 and store at 4°C. Stabilize at room temperature for 30 minutes before use.

Diluent Reagent (PBS): If using Thermo’s recommended PBS, dilute the powder to 1L of H2O and check the pH of 7.4, and store at 4°C. Stabilize at room temperature for 30 minutes before use.

Blocking Buffer: PBS + 1% Bovine Albumin (BSA). Using the PBS prepared above, dilute 0.25g of BSA in 25ml of PBS (enough for one plate). Prepare only at the time of use and do not store.

Wash Buffer: PBS + 0.1% Tween 20. To prepare 1L, add 1 sachet of PBS (Thermo) in 1L of H2O. Dissolve well and check pH 7.4. Add 1mL of Tween 20 and mix. Store at 4°C. Stabilize at room temperature for 30 minutes before use.

Substrate / chromogen (TMB): Tetramethylbenzidine for ELISA (ready-to-use solution). Store at 4°C. Stabilize at room temperature for 30 minutes before use.

STOP solution: 1N HCl (use 25mL/plate). To prepare 200mL from concentrated HCl (37%, 12N), dilute 16.7 ml of 37% HCl in 183.3 ml of H2O. Store at room temperature.

Solution of SARS-Cov-2 SPIKE antigens: resuspend in PBS, at 1.0mg/ml. Store 10uL aliquots at −80°C, avoid re-freezing. Each 10uL aliquot is enough to make a 96-well plate.

Secondary antibody: Anti-human IgG HRP-conjugated. Use 1:30,000 dilution. Prepare 10uL aliquots, store at −20°C and avoid re-freezing.

### Experimental Protocol

1. Add 10uL of the SARS-Cov-2 Antigen Solution to 10mL of the Sensitization Buffer. Pipete 100uL of the sensitization solution per well. Incubate overnight at 4°C.
2. Wash the plate with 200ul of Wash Buffer per well. Repeat twice. Discard the last washing before the next step.
3. Add 200 ul of the Blocking Buffer per well. Incubate at 37 ° C for 1 hour. Washing.
4. Wash the plate with 200ul of Wash Buffer per well. Repeat twice. Discard the last washing before the next step.
5. Load samples to the test plate. Samples should be diluted 1XPBS. Serum samples may be diluted 1:100. It’s desirable to have a negative control (negative serum for Sars-Cov-2 antibodies) and a positive control in every testing. Incubate 2 hours at 37°C.
6. Wash the plate with 200ul of Wash Buffer per well. Repeat twice. Discard the last washing before the next step.
7. Dilute HRP-anti human 1:30000 in PBS. Add 100ul per well, incubate 1 hour at 37°C.
8. Wash the plate with 200ul of Wash Buffer per well. Repeat twice. Discard the last washing before the next step.
9. Add 100ul of TMB substrate per well. Wrap plate in aluminum foil, incubate 3 minutes at room temperature. Add 100ul of STOP solution per well.
10. Read absorbance at 450nm.

## Data Availability

Additional data my be provided upopn request

## AKNOWLEDGMENT

We thanks Dr. Leda Castilho, from Universidade Federal do Rio de Janeiro (UFRJ) for kindly providing the SPIKE protein. We thanks LAM-LNBio, LEC-LNBio and LVV-LNBio to facilitate experiments for assay development. We thanks MCTi, FINEP – Rede virus, FAPESP 2019/04458-8 for financial support.

